# 3 Year Outcomes of Mitral Valve-in-Valve Therapy Using Balloon Expandable Transcatheter Valves in the United States

**DOI:** 10.1101/2024.09.20.24314103

**Authors:** Mackram F. Eleid, Amar Krishnaswamy, Samir Kapadia, Pradeep Yadav, Vivek Rajagopal, Raj Makkar, Curtiss Stinis, Stanley Chetcuti, Andrew Morse, Tiberio Frisoli, Antonio H. Frangieh, Amr E. Abbas, Brian Whisenant, William W. O’Neill, Mayra E. Guerrero, Evelio Rodriguez, Susheel Kodali, Gorav Ailawadi, Charanjit S. Rihal

## Abstract

Mitral valve-in-valve (MViV) is a safe and effective therapy for severe bioprosthetic mitral degeneration associated with high one-year survival rates. Longer-term survival and reintervention rates are not well defined.

**Methods:** Retrospective analysis of patients undergoing transseptal MViV with SAPIEN 3 valve family for failed surgical bioprostheses from June 2015 to March 2024 in the Transcatheter Valve Therapy Registry with Centers for Medicare and Medicaid Services data linkage was performed. The primary outcomes were all-cause mortality, stroke, and reintervention. Kaplan Meier and Cox proportional hazards analysis of outcomes was performed according to Society of Thoracic Surgeons (STS) score and procedure status.

**Results:** 5,971 patients (72.9 ± 11.4 years, 57.9% female) underwent MViV. Low (<4), intermediate (4-8), and high (>8) STS scores were present in 1310 (23.5%), 1960 (35.1%) and 2315 (41.5%) patients, respectively. 30-day mortality occurred in 0.9% of patients with low STS scores; 2.7%, intermediate; and 7.0%, high (p<0.001). Predicted mortality at 3 years was proportional to STS score at 15.8%, 23.3%, and 44.5% (p<0.001 for all comparisons). Stroke rates at 3 years were comparable except between low and high STS groups (7.6% vs. 11.4%, p=0.002). Mitral valve reintervention during 3 years of follow-up was similarly uncommon in all groups at 3.8, 3.0, and 2.8% (p=0.71), respectively. Elective procedures were associated with lower 3-year mortality compared to non-elective procedures (HR 0.51, [95% CI: 0.45, 0.58], p<0.001). Strongest predictors of 3-year mortality included current hemodialysis, cardiogenic shock on presentation, and immunocompromised state.

**Conclusion:** Three-year survival after MViV is favorable in low and intermediate STS scores and elective procedures, whereas survival was significantly lower in high STS scores and non-elective procedures. These findings emphasize the importance of early identification and treatment of patients who may benefit from MViV. Reintervention rates at 3 years are low regardless of STS score.

**Clinical Perspectives:** *What is new?:*

- Three-year survival after transseptal mitral valve-in-valve (MViV) is favorable in patients with low and intermediate Society of Thoracic Surgeons (STS) risk scores
- High STS risk score and non-elective procedure status are associated with higher 3-year mortality
- Reintervention rates after MViV are low at 3 years regardless of STS score

*What are the clinical implications?:*

- MViV is an effective therapy associated with favorable mid-term outcomes in carefully selected patients
- Strategies to treat patients before advanced comorbidities develop are needed to optimize longer-term outcomes following MViV

## Introduction

Over the last decade, transseptal mitral valve-in-valve (MViV) has developed into a first-line therapy for patients with degenerated mitral bioprostheses as a less invasive alternative for patients at increased risk for repeat cardiac surgery.^1,2^ Multiple studies have demonstrated the favorable short-term safety and efficacy of MViV, with high 30-day and 1-year survival rates.^3–5^ However, only limited data is available regarding mid-term outcomes of transseptal MViV.^6–8^ Given the inherently smaller effective orifice resulting from the implantation of a balloon-expandable valve within a degenerated bioprosthesis with associated mildly increased mean Doppler gradients, questions have been raised regarding the long-term durability and outcomes of this groundbreaking minimally invasive therapy. These questions deepen as MViV therapy is applied to lower surgical risk populations who could potentially undergo either redo surgery or MViV.^2^ Mid-term outcome data using the newest generation SAPIEN 3 and SAPIEN 3 ultra balloon-expandable valves in the United States has not yet been reported but could further inform the role of MViV in the treatment of the spectrum of patients presenting with mitral bioprosthesis degeneration. To better understand mid-term outcomes of this novel therapy, the goal of this study was to evaluate survival, stroke, and reintervention following MViV in a large population at 3 years of follow-up using the Society of Thoracic Surgeons (STS)/American College of Cardiology (ACC) Transcatheter Valve Therapy (TVT) Registry.

## Methods

### Study Population

Patients undergoing transseptal MViV in the United States from June 2015 through March 2024 reported to the STS/ACC TVT Registry were included in this study. The TVT Registry has been approved by a central institutional review board (Advarra) with a waiver of informed consent granted by the Duke University School of Medicine institutional review board under the Common Rule 45 CFR 46.3. This article conforms to the Strengthening of Reporting of Observational Studies in Epidemiology cohort guidelines.

The study inclusion criteria included patients undergoing transseptal MViV for degenerated bioprosthetic mitral valves. Patients undergoing transapical, other alternative access MViV, or mitral valve-in-ring or native mitral valve anatomy were excluded.

### Procedure

Transseptal MViV procedural techniques have been previously described and include transvenous access with the creation of a balloon atrial septostomy to facilitate delivery of the balloon-expandable SAPIEN 3 valve over a curved guidewire positioned in the left ventricular apex. The valve is carefully positioned under fluoroscopic and echocardiographic guidance and deployed using rapid ventricular pacing, with the failed bioprosthetic valve as an anchor. Following implantation, the delivery system is removed, and the atrial septal defect is assessed and closed with an atrial septal defect closure device if deemed necessary.

### Outcome definitions

The primary outcomes of interest were all-cause mortality, stroke and repeat mitral intervention at 3 years. Secondary outcomes included New York Heart Association (NYHA) class and quality of life defined by the Kansas City Cardiomyopathy Questionnaire (KCCQ) at 1 year. The ratio of observed to expected mortality was reported with expected mortality defined as the STS risk score for surgical mortality.

### Statistical Analysis

Continuous variables were presented as mean (standard deviation [SD]) or median (interquartile range [IQR]) and were compared between groups using the two-sample t-test or Wilcoxon rank sum test. Categorical variables were presented as frequencies and percentages and were compared using the χ^2^ or Fisher’s exact test. The 30-day, 1-year, and 3-year adverse event rates were based on Kaplan–Meier estimates, and all comparisons were made using the log-rank test.

A multivariable Cox regression model was performed to identify independent predictors of 3-year mortality. Baseline comorbidities with p-value <0.1 in the univariable analysis were selected as candidate covariates. Thirty-one covariates were used for adjustment: age, race, procedure status, atrial fibrillation/flutter, porcelain aorta, hostile chest, carotid stenosis, number of diseased vessels, prior percutaneous coronary intervention (PCI), prior coronary artery bypass graft (CABG) surgery, prior myocardial infarction, previous implantable cardioverter-defibrillator (ICD), permanent pacemaker, currently on dialysis, chronic lung disease, home oxygen, hemoglobin, creatinine, glomerular filtration rate (GFR), diabetes mellitus, peripheral arterial disease, ≥ moderate aortic regurgitation, left ventricular ejection fraction (LVEF) <50, cardiac index < 2.2, ≥ moderate mitral regurgitation, ≥ moderate tricuspid regurgitation, immunocompromised, cardiogenic shock within 24 hours, heart failure hospitalization within past year, NYHA III/IV, and KCCQ-OS. Proportional-hazards assumption was confirmed through testing based on Kolmogorov-type Supremum Test. A Cox regression model was used with stepwise selection, which consisted of entering the model covariates with p ≤ 0.10 and removing covariates with p > 0.10.

To perform 3-year survival, stroke, and reintervention analyses, the STS/ACC TVT Registry was linked to the United States Centers for Medicare and Medicaid Services (CMS) claims data using probabilistic matching with patient birth date, gender and TAVR procedure date. Patients eligible for linkage were all over 65 years with Medicare coverage enrolled in the Medicare Parts A and B fee-for-service program. Multiple matches were removed, and only one unique case was used for the analysis.

All p-values were 2-sided, p < 0.05 was considered significant for all tests, and no adjustment for multiple testing was undertaken. All statistical analyses were performed using SAS version 9.4 (SAS Institute Inc., Cary, North Carolina).

## Results

### Baseline Characteristics

A total of 5,971 patients (mean age 72.9 ± 11.4 years, 57.9% female, 82.5% Caucasian, 10.1% African American) underwent transseptal MViV and were included in the study (Figure 1 and Table 1). Baseline clinical and echocardiographic characteristics are shown in Table 1. Low (<4), intermediate (4-8), and high (>8) STS scores were present in 1310 (23.5%), 1960 (35.1%), and 2315 patients (41.5%), respectively (Figure 1 and Supplemental Table 1). The mean STS scores in each group were as follows: 2.7 ± 0.9 in low-risk, 5.9 ± 1.2 in intermediate-risk and 15.5 ± 9.4 in the high-risk group. Lower-risk patients were significantly younger than intermediate-risk and high-risk (66.5 ± 12.7 versus 73.8 ± 9.5 versus 76.2 ± 10 years, respectively, p<0.0001). In the overall population, 74.2% of patients had one prior cardiac surgery, 19.1% had two prior cardiac surgeries, and 3.5% had three or more. Prior stroke was present in 18.3% of the overall population, hypertension in 84.1%, atrial fibrillation/flutter in 73%, and diabetes mellitus in 27.5%. Renal failure requiring dialysis was more common in the high-risk group (8.6%) compared to the low-risk (1.3%) and intermediate-risk (2.7%) groups (p<0.0001). Home oxygen use was more common in the high-risk group (21%) compared to the low-risk (4.1%) and intermediate-risk (10.3%) groups (p<0.0001). Prior endocarditis was more common in the low-risk group. Patients in higher-risk groups were more likely to present with NYHA class III/IV symptoms (88.3% versus 72.3% in low-risk) and more likely to present in cardiogenic shock (present in 7% of high-risk patients at baseline compared to 1% in low-risk). Baseline quality of life measured by the KCCQ was worse in the higher-risk groups (p<0.0001). Concomitant valvular heart disease, including aortic regurgitation, aortic stenosis, and tricuspid regurgitation, were all more common in higher-risk patients (p<0.05 for all).

**Figure 1:**
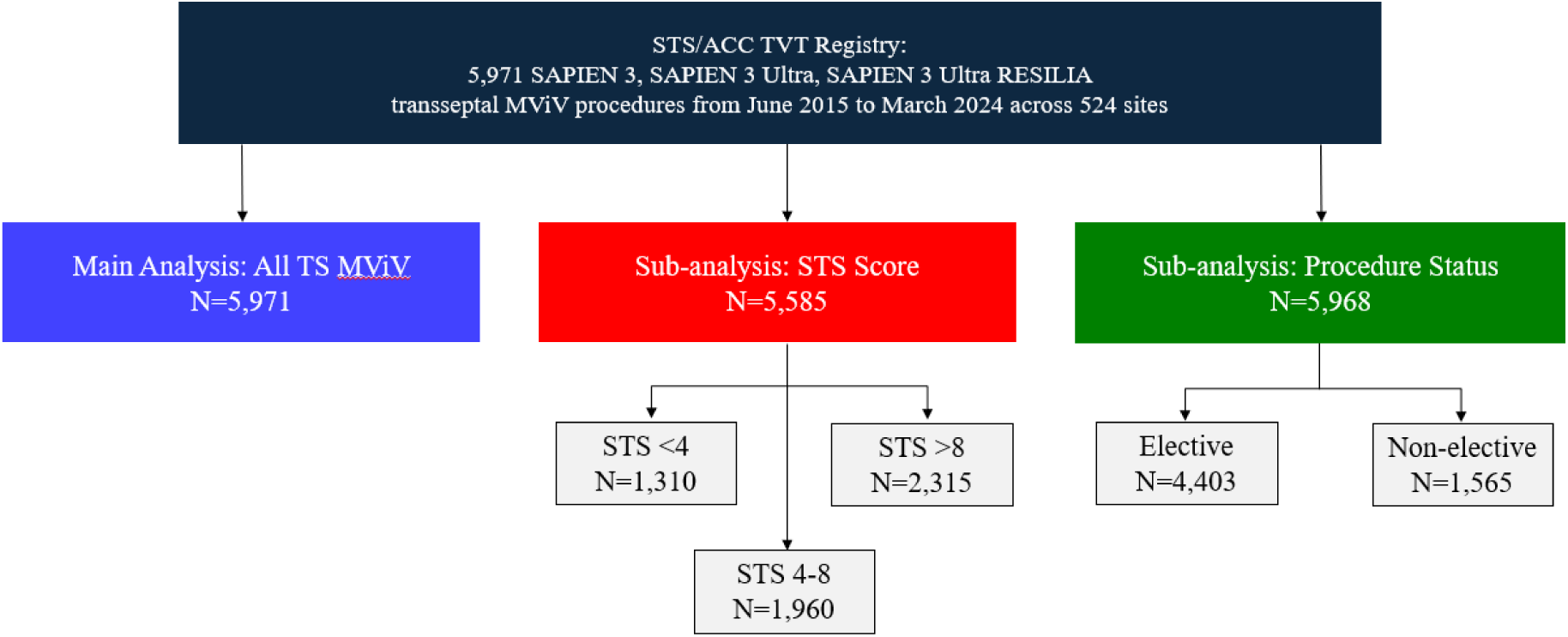
Study flow chart: Consecutive patients undergoing transseptal mitral valve in valve (MViV) from June 2015 to March 2024 were evaluated as an overall cohort (5,971) as well as compared according to Society of Thoracic Surgeons risk score status (N=5,585) and according to elective versus non-elective procedure status (N=5,968). ACC indicates American College of Cardiology; MViV, mitral valve-in-valve; STS, Society of Thoracic Surgeons; TS, transseptal; and TVT, transcatheter valve therapy.

**Table 1.** Baseline Characteristics.

|  | MViV (N=5971) |
| --- | --- |
| Clinical characteristic |  |
| Age (years) | 72.9 ± 11.4 (5971) |
| Female | 57.9% (3457/5971) |
| STS score (%) | 9.1 ± 8.2 (5585) |
| BMI (kg/m<sup>2</sup>) | 27.6 ± 13.3 (5956) |
| Permanent pacemaker | 26.4% (1571/5962) |
| Previous ICD | 10.6% (632/5960) |
| Prior MI | 17.1% (1019/5965) |
| Prior PCI | 15.8% (945/5965) |
| Prior CABG | 29.7% (1769/5964) |
| Prior aortic valve procedure | 23.9% (1428/5969) |
| Prior stroke | 18.3% (1091/5969) |
| Prior TIA | 9.7% (576/5970) |
| Hypertension | 84.1% (5019/5969) |
| Immunocompromised | 7.4% (407/5534) |
| Endocarditis | 14.0% (835/5966) |
| Diabetes mellitus | 27.5% (1640/5965) |
| Peripheral arterial disease | 13.9% (830/5965) |
| GFR | 56.6 ± 25.0 (5952) |
| Currently on dialysis | 4.9% (292/5963) |
| Hostile chest | 12.4% (741/5969) |
| Carotid stenosis | 9.7% (486/5020) |
| Heart failure hospitalization within past year | 58.1% (2896/4986) |
| Cardiogenic shock within 24 hours | 4.1% (245/5967) |
| Porcelain aorta | 0.4% (25/5966) |
| Atrial fibrillation/flutter | 73.0% (4357/5969) |
| Chronic lung disease | 37.6% (2233/5944) |
| Home oxygen | 13.2% (788/5964) |
| Positive Inotropes | 9.2% (545/5941) |
| BNP | 779.3 ± 1037.7 (2439) |
| NT proBNP | 5088.7 ± 7757.0 (2019) |
| Hemoglobin | 11.6 ± 2.1 (5952) |
| Operator reason for procedure |  |
| Inoperable/Extreme risk | 17.2% (1014/5896) |
| High risk | 61.4% (3619/5896) |
| Intermediate risk | 16.0% (942/5896) |
| Low risk | 5.4% (321/5896) |
| NYHA class III/IV | 81.3% (4768/5864) |
| KCCQ overall summary score | 37.2 ± 24.0 (5052) |
| Echocardiographic characteristic |  |
| Left main stenosis ≥50% | 4.1% (207/5015) |
| Mitral annular calcification | 42.4% (919/2166) |
| MV area (cm<sup>2</sup>) | 1.4 ± 1.0 (4365) |
| MV mean gradient (mmHg) | 13.1 ± 6.2 (5698) |
| LVEF (%) | 55.1 ± 11.7 (5914) |
| ≥ Moderate mitral regurgitation | 53.2% (3153/5924) |
| Mitral stenosis | 75.7% (4409/5828) |
| ≥ Moderate aortic regurgitation | 9.6% (565/5916) |
| Aortic stenosis | 16.7% (986/5900) |

|  | **MViV (N=5971)** |
| --- | --- |
| ≥ Moderate tricuspid regurgitation | 57.0% (3388/5945) |
| Pulmonary capillary wedge pressure (mmHg) | 26.2 ± 8.5 (2870) |
| Pulmonary artery pressure (mmHg) | 40.5 ± 12.3 (2976) |
| Right atrial pressure/CVP (mmHg) | 11.9 ± 6.6 (3187) |
| Pulmonary vascular resistance | 306.5 ± 289.8 (2513) |

Values are reported as % (n/N), mean ± standard deviation (n). BMI, body mass index; BNP, b-type natriuretic peptide; NT pro-BNP, n-terminal pro b-type natriuretic peptide; CABG, coronary artery bypass graft; cm, centimeters; CVP; central venous pressure; GFR, glomerular filtration rate; ICD, implantable cardioverter-defibrillator; KCCQ, Kansas City Cardiomyopathy Questionnaire; kg, kilograms; LAD, left anterior descending; LVEF, left ventricular ejection fraction; m, meters; MI, myocardial infarction; mmHg, millimeters of mercury; MV, mitral valve; MViV, mitral valve-in-valve; NYHA, New York Heart Association; PAP, pulmonary artery pressure; PCI, percutaneous coronary intervention; PCWP, pulmonary capillary wedge pressure; PVR, pulmonary vascular resistance; STS, Society of Thoracic Surgeons; and TIA, transient ischemic attack.

### Procedural Characteristics and In-Hospital Outcomes

All procedures were performed by the transfemoral transseptal approach with no procedures performed via transapical or direct transatrial approaches. General anesthesia was used in 93.7% of procedures (Table 2). Bioprosthetic valve fracture was attempted in a total of 260 (8.5%) cases, and post-implant balloon inflation was performed in 1263 (22.7%) of cases. Successful device implantation occurred in 97.2% of patients in the overall group, with slightly higher success rates observed in the low-risk and intermediate-risk groups compared to high-risk (97.4% and 98.0% versus 96.4%, respectively, p=0.006; Supplemental Table 1). Left ventricular outflow tract obstruction occurred in 14 (0.2%) patients, atrial septal defect closure was performed in 528 (8.8%), and procedural mortality occurred in 28 patients (0.5%).

**Table 2.** Procedural and In-Hospital Outcomes.

|  | MViV (N=5971) |
| --- | --- |
| Device implanted successfully | 97.2% (5802/5971) |
| MVARC technical success | 96.9% (5788/5971) |
| Procedure status |  |
| Elective | 73.8% (4403/5968) |
| Urgent | 24.2% (1445/5968) |
| Emergency | 1.5% (92/5968) |
| Salvage | 0.5% (28/5968) |
| Anesthesia type |  |
| General anesthesia | 93.7% (5590/5966) |
| Moderate sedation | 6.0% (359/5966) |
| Conversion to open heart surgery | 0.6% (36/5970) |
| Total procedure time (min) | 102.0 ± 55.9 (5966) |
| Fluoroscopy time (min) | 32.5 ± 21.4 (5377) |
| Contrast volume (ml) | 16.9 ± 39.6 (4779) |
| THV type |  |
| S3 | 64.9% (3875/5971) |
| S3 Ultra | 23.5% (1402/5971) |
| S3 Ultra Resilia | 11.6% (694/5971) |
| THV size |  |
| 20 mm | 0.1% (5/5968) |
| 23 mm | 6.6% (393/5968) |
| 26 mm | 42.3% (2522/5968) |
| 29 mm | 51.1% (3048/5968) |
| Bioprosthetic valve fracture attempted | 8.5% (260/3071) |
| Mechanical support | 3.3% (199/5965) |

|  | **MViV (N=5971)** |
| --- | --- |
| Procedure aborted | 0.3% (15/5971) |
| Transseptal complication | 0.6% (35/5971) |
| Device embolization | 0.2% (14/5971) |
| Device migration | 0.3% (16/5971) |
| Procedural mortality | 0.5% (28/5971) |
| Length of stay (days) | 2.0 [1.0, 4.0] |
| ICU length of stay (hours) | 23.0 [0.0, 37.0] |
| Discharged w/ anticoagulants | 82.9% (4730/5708) |
| Discharged w/ antiplatelets | 72.2% (4122/5712) |
| Discharged home | 86.8% (5185/5971) |

\*MVARC technical success was defined as at exit from the hybrid suite, patient is alive with successful access, delivery, and retrieval of the device delivery system, successful deployment and correct position of the first intended device, and freedom from emergency surgery or reintervention associated with the device or access procedure.
Values are reported as % (n/N), median [IQR], or mean ± standard deviation (n). Abbreviations: BVF, bioprosthetic valve fracture; hrs, hours; ICU, intensive care unit; min, minutes; MVARC, Mitral Valve Academic Research Consortium; MViV, mitral valve-in-valve; and THV, transcatheter heart valve.

In-hospital mortality occurred in 3.1% of the overall population and was more frequent in the high-risk group (5.3% versus 0.5% and 1.8% in the low-risk and intermediate-risk groups; Table 4 and Supplemental Table 4). In-hospital stroke rates were low in all three groups (0.4%, 1.0%, and 1.3% in low, intermediate, and high-risk groups, respectively, p=0.03). Hospital length of stay was longer for higher-risk groups (p<0.0001), and 86.8% of the overall group were discharged to home. Regarding medical therapy, 82.9% were prescribed anticoagulation, and 72.2% were prescribed antiplatelet therapy at hospital discharge.

### One-Year Outcomes

At one year, NYHA class III/IV improved compared to baseline for the overall group (81.3% to 13.7%; Tables 1 and 4). High-risk and intermediate-risk patients were more likely to have class II symptoms at 1 year compared to the low-risk group, but all three groups similarly had a low prevalence of class III or IV symptoms at one year (low-risk: 11.8%, intermediate-risk: 13.1%, and high-risk: 15.4%; Supplemental Table 4). Quality of life as measured by the KCCQ improved by 39.3 ± 28.3 points compared to baseline in the overall group, improvement was observed across all risk groups (low-risk: 36.7 ± 27.9, intermediate-risk: 37.9 ± 27.7, and high-risk: 42.4 ± 29.0, p=0.002). All-cause predicted mortality at one year was 14.1% in the overall group, 5.5% in low-risk, 10.4% in intermediate-risk, and 21.3% in high-risk groups (p<0.0001).

**Table 3.** Discharge, 30-Day, and One-Year Echocardiographic Outcomes.

|  | MViV (N=5971) |
| --- | --- |
| Discharge |  |
| MV area (cm<sup>2</sup>) | 2.0 ± 1.0 (2836) |
| MV mean gradient (mmHg) | 6.5 ± 2.8 (5401) |
| ≥ Moderate PVL | 0.3% (12/4647) |
| ≥ Moderate mitral regurgitation | 0.6% (35/5496) |
| ≥ Moderate tricuspid regurgitation | 37.7% (1154/3061) |
| 30 Days |  |
| MV area (cm<sup>2</sup>) | 1.9 ± 1.0 (2255) |
| MV mean gradient (mmHg) | 7.5 ± 3.0 (4028) |
| LVEF (%) | 54.0 ± 11.4 (4106) |
| ≥ Moderate PVL | 0.2% (7/3405) |
| ≥ Moderate mitral regurgitation | 0.7% (29/4096) |
| ≥ Moderate tricuspid regurgitation | 38.9% (1583/4065) |
| 1 Year |  |
| MV area (cm<sup>2</sup>) | 1.9 ± 1.0 (995) |
| MV mean gradient (mmHg) | 7.5 ± 3.2 (1906) |
| LVEF (%) | 54.1 ± 11.3 (1995) |
| ≥ Moderate PVL | 0.4% (6/1554) |
| ≥ Moderate mitral regurgitation | 0.9% (17/1950) |
| ≥ Moderate tricuspid regurgitation | 33.8% (661/1957) |

Values are reported as % (n/N), mean ± standard deviation (n). Abbreviations: cm, centimeters;
LVEF, left ventricular ejection fraction; mmHg, millimeters of mercury; MV, mitral valve;
MViV, mitral valve-in-valve; and PVL, paravalvular leak.

**Table 4.** In-hospital, 30-Day, and One-Year Clinical Outcomes.

|  | MViV (N=5971) |
| --- | --- |
| **In Hospital Outcome** |  |
| All-cause mortality | 3.1% (187/5971) |
| Cardiac death | 1.8% (110/5971) |
| Mitral valve re-intervention | 0.4% (22/5971) |
| Major vascular complication | 1.3% (76/5971) |
| Life threatening bleeding | 1.3% (78/5971) |
| New requirement for dialysis | 1.0% (59/5971) |
| Myocardial infarction | 0.1% (8/5971) |
| New pacemaker w/o baseline pacemaker | 1.2% (51/4400) |
| New onset atrial fibrillation | 1.4% (49/3563) |
| Stroke | 1.0% (61/5971) |
| TIA | 0.1% (5/5971) |
| LVOT obstruction | 0.5% (27/5971) |
| Device thrombosis | 0.2% (11/5971) |
| Cardiac perforation | 0.9% (56/5971) |
| ASD closure | 8.8% (528/5971) |
| Any readmission | 0.2% (14/5971) |
| Cardiac readmission | 0.1% (7/5971) |
| Heart failure readmission | 0.0% (2/5971) |
| **30 Days Outcome** |  |
| All-cause mortality | 4.2% (244) |
| Cardiac death | 2.2% (126) |
| Mitral valve re-intervention | 0.5% (30) |
| Major vascular complication | 1.5% (85) |
| Life threatening bleeding | 1.4% (78) |
| New requirement for dialysis | 1.2% (66) |
| Myocardial infarction | 0.3% (16) |
| New pacemaker w/o baseline pacemaker | 1.5% (61) |
| New onset atrial fibrillation | 1.7% (57) |
| Stroke | 1.6% (96) |
| TIA | 0.2% (10) |
| LVOT obstruction | 0.5% (27) |
| Device thrombosis | 0.2% (13) |
| ASD closure | 9.0% (533) |
| Any readmission | 9.1% (501) |
| Cardiac readmission | 1.7% (95) |
| Heart failure readmission | 2.2% (122) |
| NYHA class |  |
| Class I | 45.3% (1727/3812) |
| Class II | 39.9% (1519/3812) |
| Class III | 13.2% (502/3812) |
| Class IV | 1.7% (64/3812) |
| KCCQ overall summary score - Change from Baseline to 30 Days | 35.4 ± 28.5 (3564) |
| 1 Year Outcome |  |
| All-cause mortality | 14.1% (690) |
| Cardiac death | 5.1% (232) |
| Mitral valve re-intervention | 1.6% (77) |
| Major vascular complication | 1.7% (95) |
| Life threatening bleeding | 2.6% (124) |
| New requirement for dialysis | 1.8% (88) |
| Myocardial infarction | 0.8% (34) |

|  | **MViV (N=5971)** |
| --- | --- |
| New pacemaker w/o baseline pacemaker | 3.0% (103) |
| New onset atrial fibrillation | 3.1% (90) |
| Stroke | 3.9% (190) |
| TIA | 0.6% (24) |
| LVOT obstruction | 0.5% (27) |
| Device thrombosis | 0.5% (23) |
| ASD closure | 9.8% (564) |
| Anticoagulant use | 70.4% (1931/2745) |
| Antiplatelet use | 59.6% (1633/2740) |
| Any readmission | 32.3% (1372) |
| Cardiac readmission | 8.7% (345) |
| Heart failure readmission | 10.5% (434) |
| NYHA class |  |
| Class I | 48.7% (959/1968) |
| Class II | 37.6% (740/1968) |
| Class III | 12.3% (241/1968) |
| Class IV | 1.4% (28/1968) |
| KCCQ overall summary score - Change from baseline to 1 year | 39.3 ± 28.3 (1773) |

Values are % (n/N) for in-hospital outcomes or Kaplan Meier estimate % (number of events) for 30-day and 1-year outcomes. Abbreviations: ASD, atrial septal defect; LVOT; left ventricular outflow tract; KCCQ, Kansas City Cardiomyopathy Questionnaire; MViV, mitral valve-in-valve; NYHA, New York Heart Association; and TIA, transient ischemic attack.

At one year, 70.4% of the overall group were prescribed an anticoagulant and 59.6% of patients were taking antiplatelet therapy. Echocardiographic mean gradients were similar between groups at 1 year (low-risk: 7.6 ± 2.9, intermediate-risk: 7.6 ± 3.2, and high-risk: 7.3 ± 3.4 mmHg, p=0.18). Calculated mitral valve area by echocardiogram was also similar between groups (low-risk: 1.9 ± 1.3, intermediate-risk: 1.8 ± 0.8, and high-risk: 1.9 ± 1.0 cm^2^, p=0.21). Mitral paravalvular leak was uncommon in the overall group (96.5% had no paravalvular leak) and similar between groups (low-risk: 96.7%, intermediate-risk: 95.6%, and high-risk: 95% with no paravalvular leak, p=0.45).

### Three-Year Clinical Outcomes

In the overall group, 3-year predicted mortality was 31.9%, stroke was 9.9%, and reintervention 3.2% (Figures 2A-C). Predicted mortality at 3 years was proportional to STS score at 15.8%, 23.3%, and 44.5% (p<0.0001 for all comparisons, Figure 2A). Stroke rates at 3 years were comparable except between low and high STS groups (7.6% vs. 11.4%, p=0.002; Figure 2B). Mitral valve reintervention during 3 years of follow-up was similarly uncommon in all groups at 3.8%, 3.0%, and 2.8% (p=0.71), respectively (Figure 2C).

**Figure 2A:**
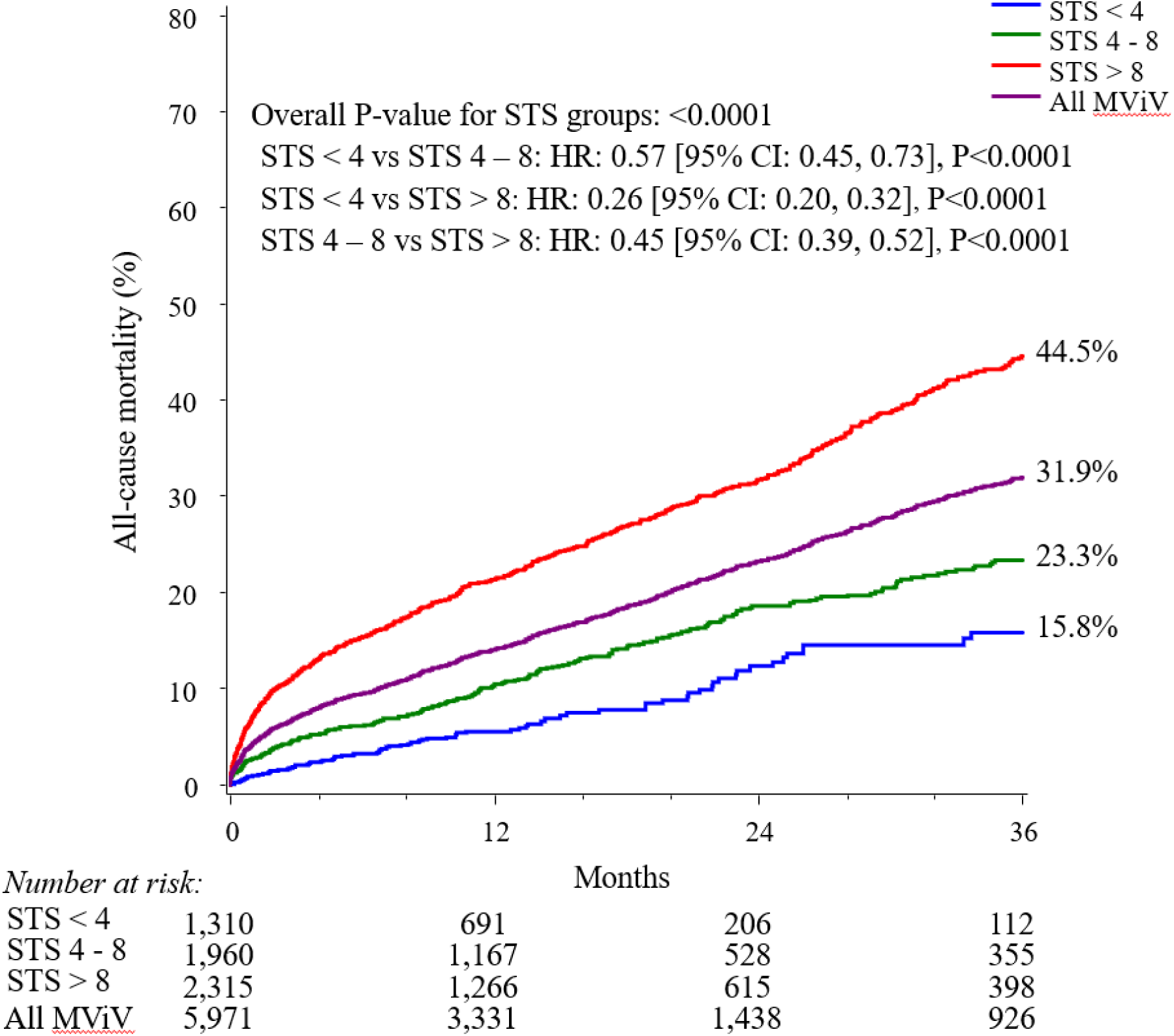
3-year outcomes in the overall cohort and According to STS Score —All-cause mortality. *All MViV group includes all transseptal MViV patients (N=5971). STS groups exclude patients with missing STS scores (N=5585). Higher STS score was associated with higher risk of mortality. CI indicates confidence interval; HR, hazard ratio; MViV, mitral valve-in-valve and STS, Society of Thoracic Surgeons.

**Figure 2B:**
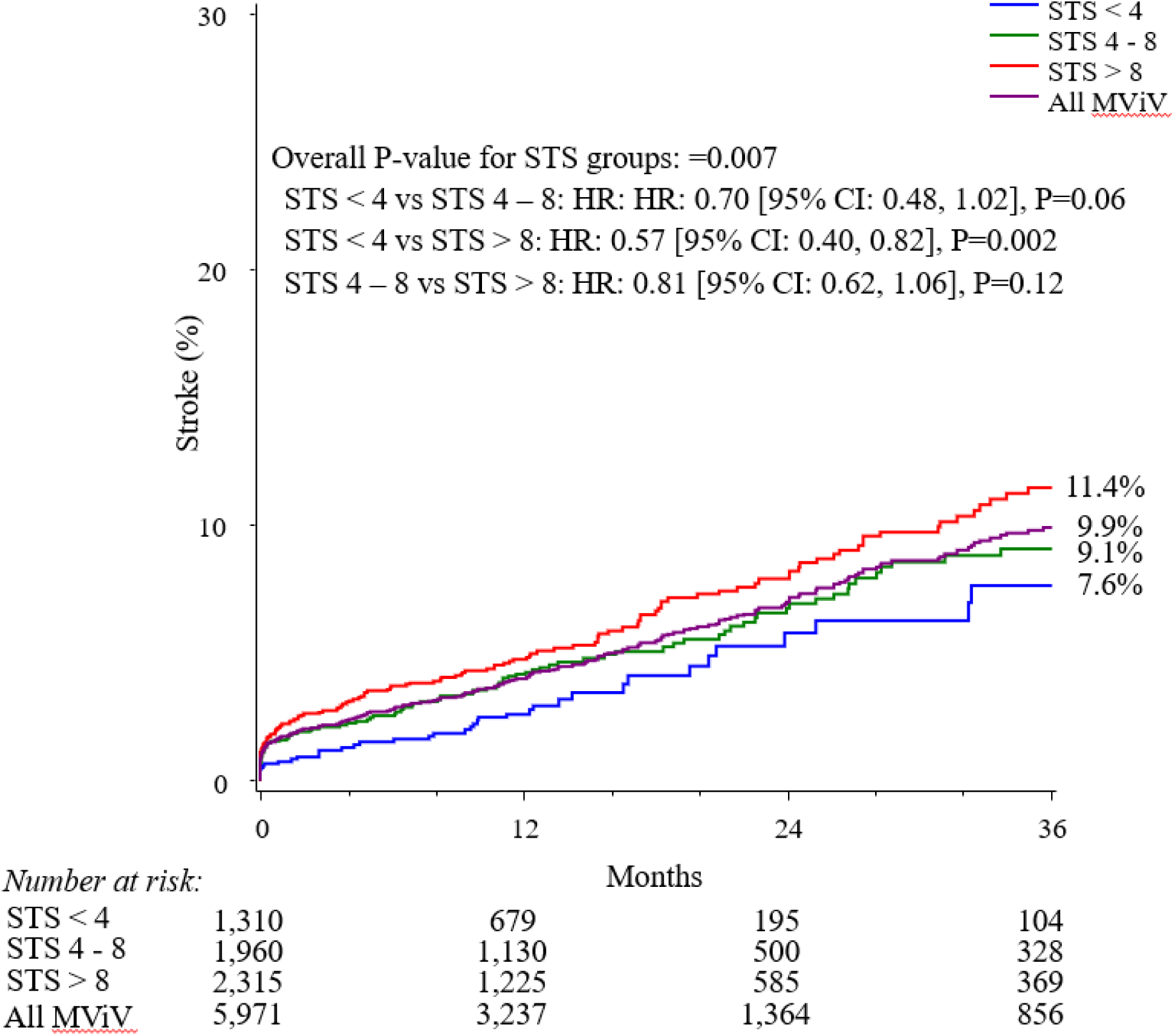
3-year outcomes in the overall cohort and According to STS Score —Stroke. *All MViV group includes all transseptal MViV patients (N=5971). STS groups exclude patients with missing STS scores (N=5585). Higher STS score was associated with higher risk of stroke. CI indicates confidence interval; HR, hazard ratio; MViV, mitral valve-in-valve, and STS, Society of Thoracic Surgeons.

**Figure 2C:**
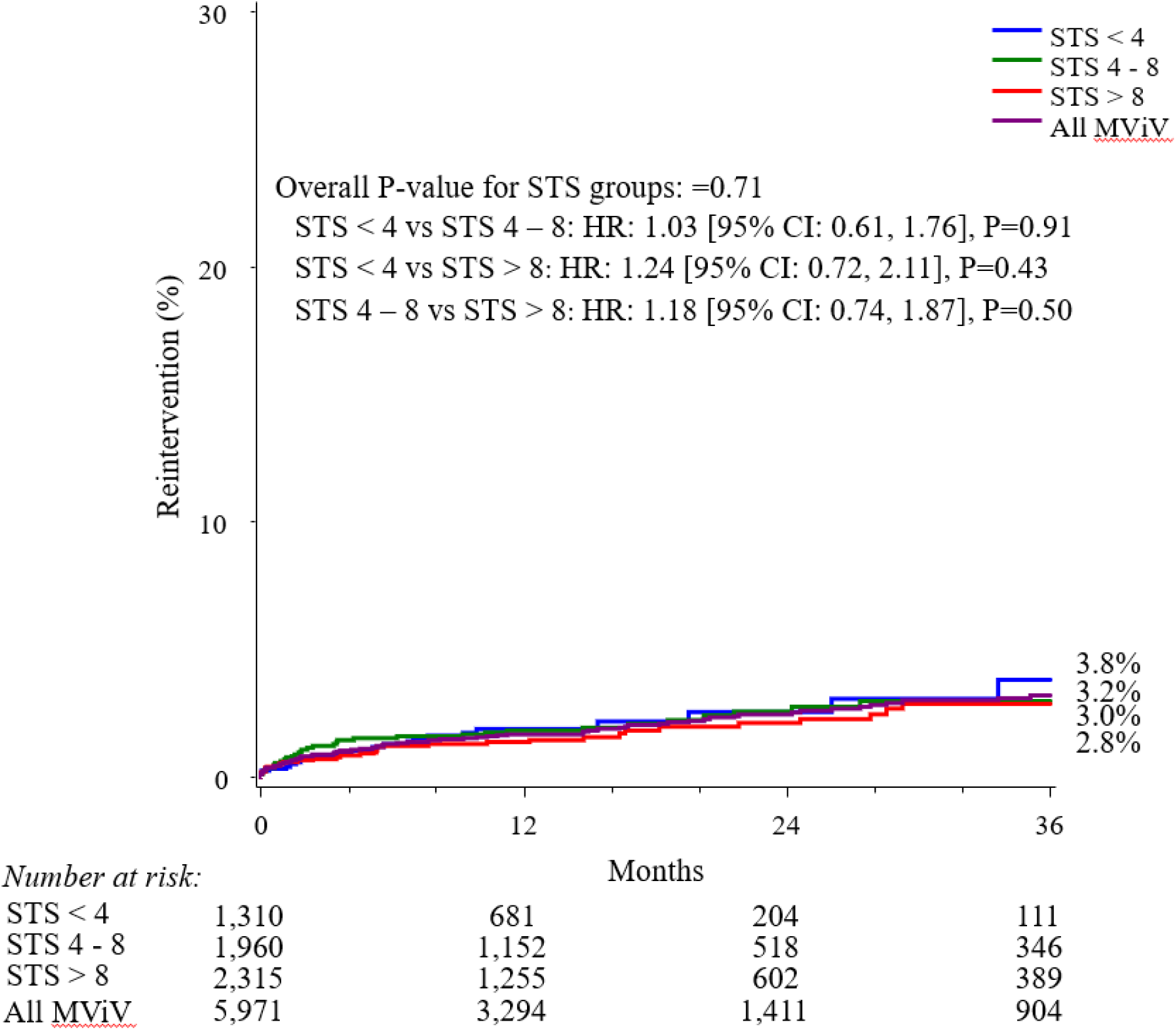
3-year outcomes in the overall cohort and According to STS Score — Reintervention. *All MViV group includes all transseptal MViV patients (N=5971). STS groups exclude patients with missing STS scores (N=5585). Higher STS score was associated with similar rates of reintervention. CI indicates confidence interval; HR, hazard ratio; MViV, mitral valve-in-valve, and STS, Society of Thoracic Surgeons.

### Outcomes According to Procedure Status

Elective procedures were associated with lower 3-year mortality compared to non-elective procedures (28.2% versus 43.3%; Figure 3A) (HR 0.51, [95% CI: 0.45, 0.58], p<0.0001). Stroke and reintervention rates were similar between elective and non-elective procedure status at 3 years (Figures 3B and 3C).

**Figure 3A:**
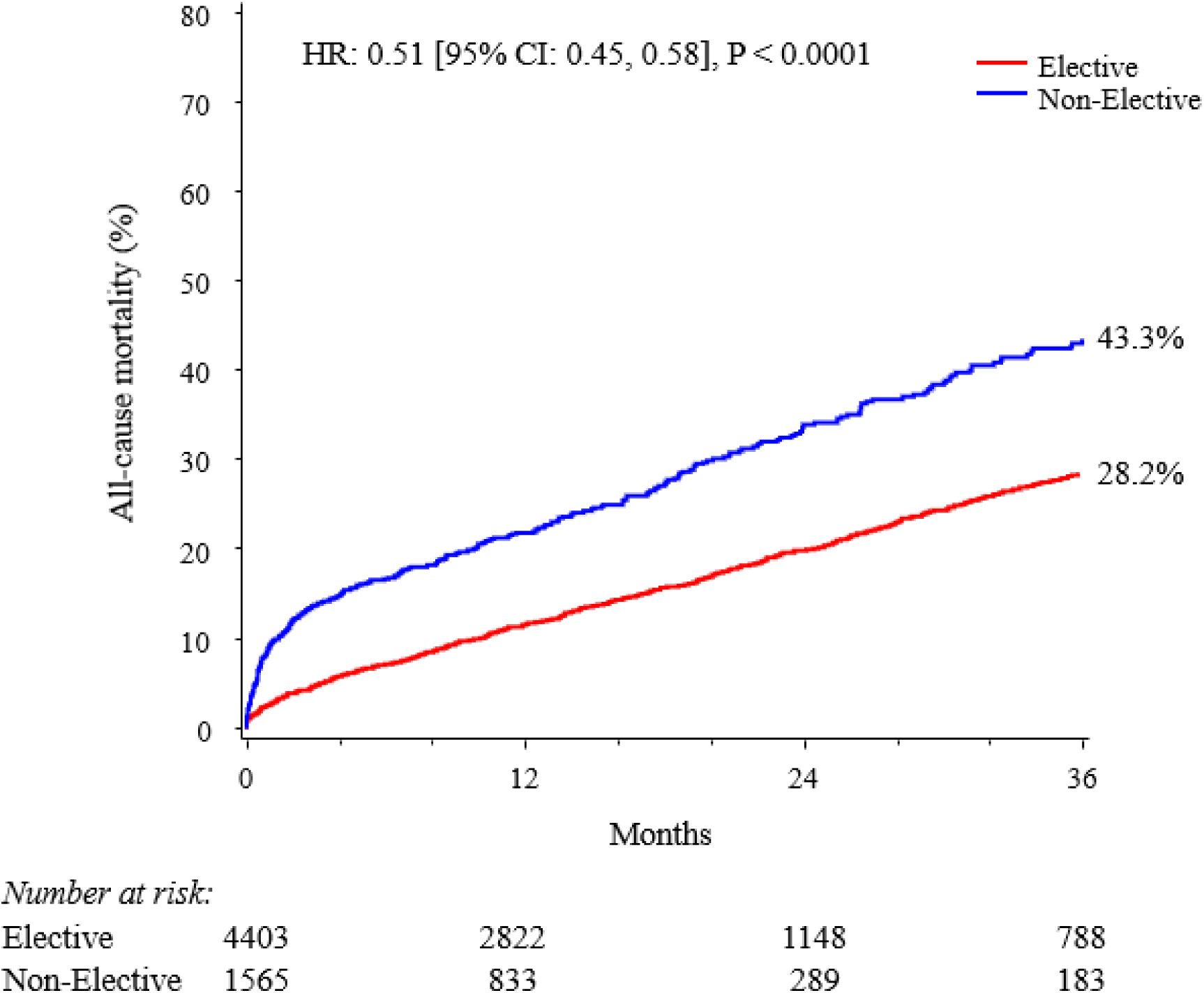
3-year outcomes according to procedure status—Mortality. Non-elective procedures were associated with higher risk of mortality. CI indicates confidence interval and HR, hazard ratio.

**Figure 3B:**
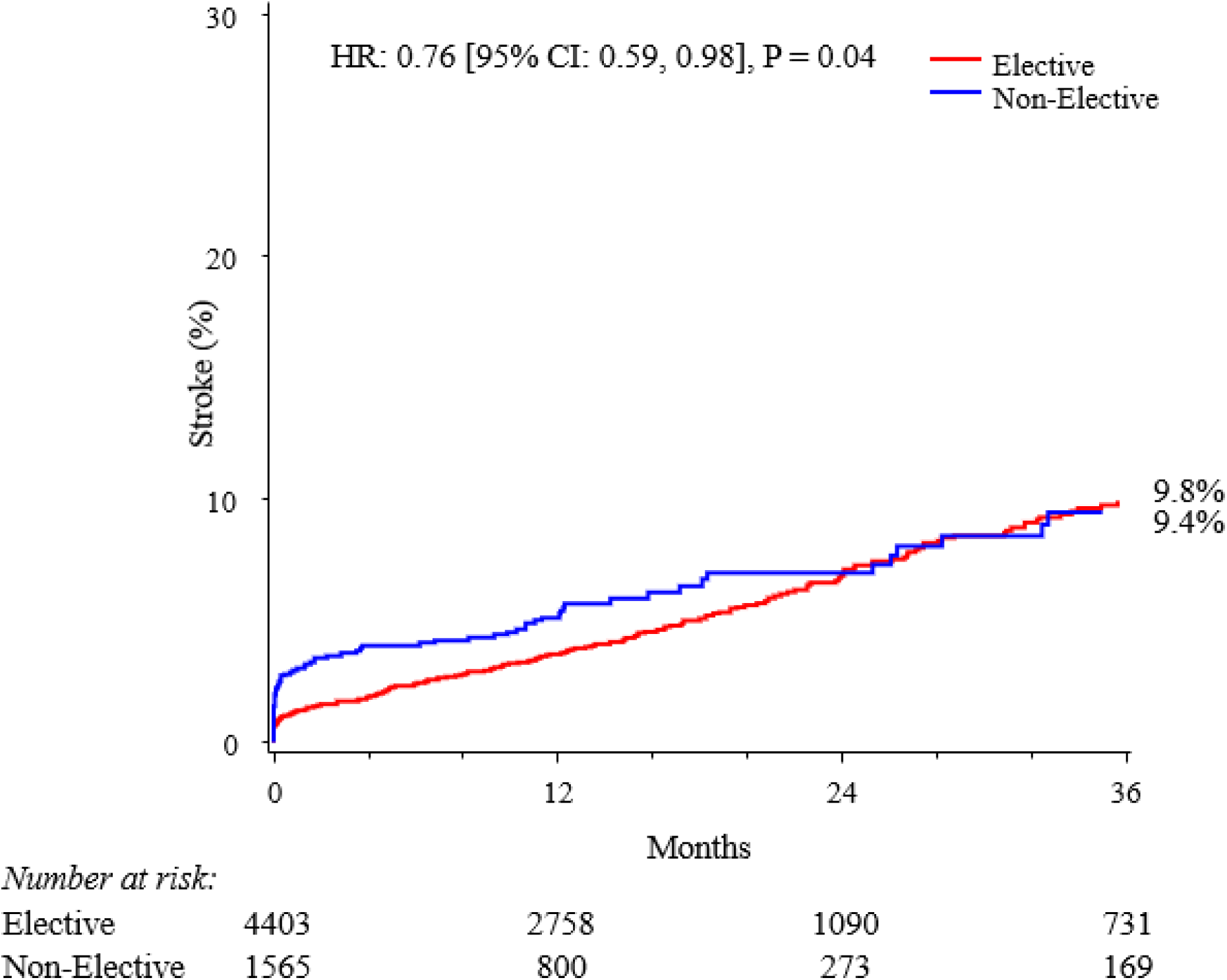
3-Year Outcomes According to Procedure Status—Stroke. Non-elective procedures were associated with higher risk of stroke. CI indicates confidence interval and HR, hazard ratio.

**Figure 3C:**
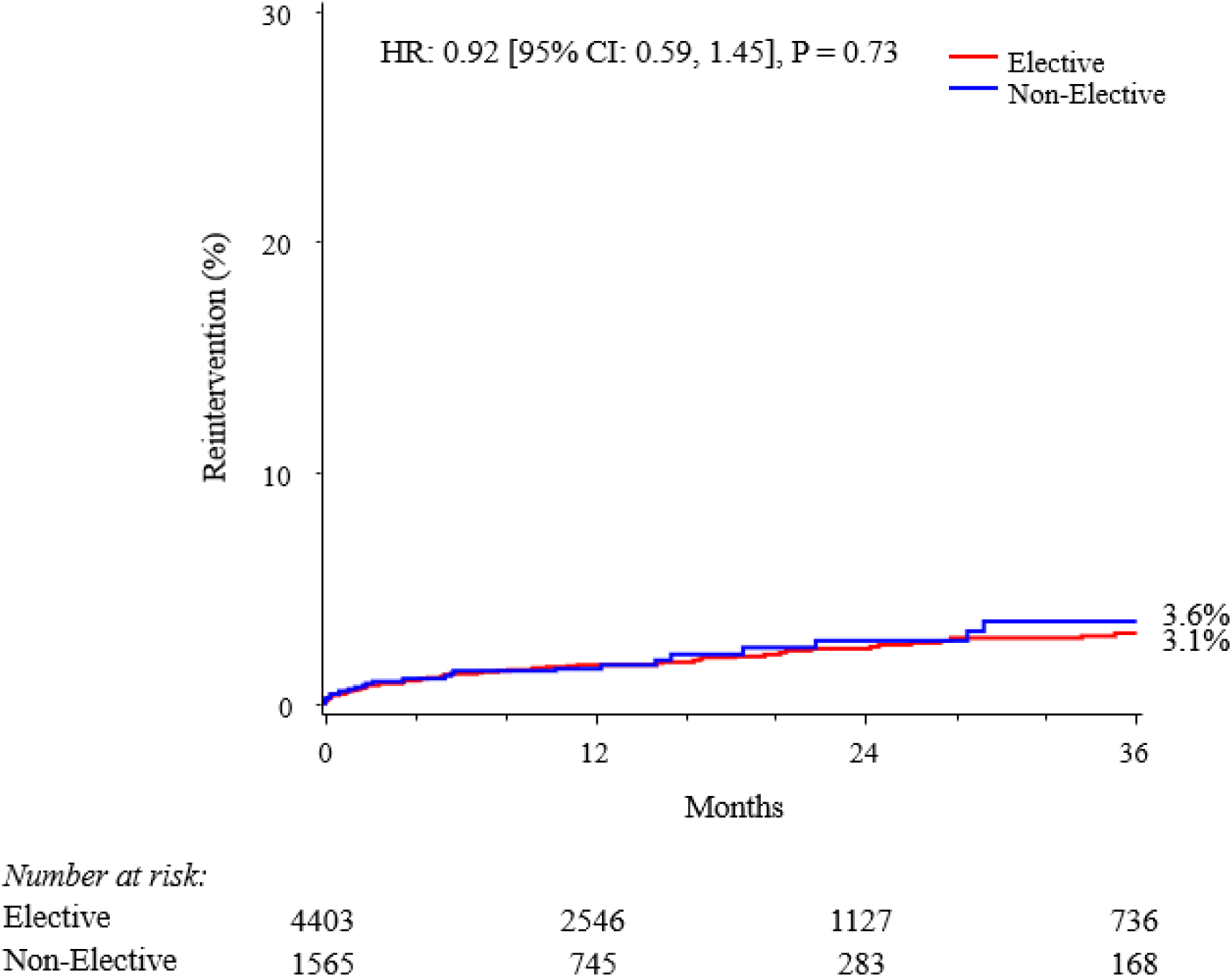
3-year outcomes according to procedure status—Reintervention. Non-elective procedures were associated with similar rates of reinterevention. CI indicates confidence interval and HR, hazard ratio. CI indicates confidence interval and HR, hazard ratio. CI indicates confidence interval and HR, hazard ratio. CI indicates confidence interval and HR, hazard ratio. CI indicates confidence interval and HR, hazard ratio.

### Predictors of Mortality

On multivariable analysis, several predictors of 3-year mortality were identified (Figure 4). Strongest predictors of 3-year mortality included current hemodialysis, cardiogenic shock on presentation, and immunocompromised state. Additional significant predictors of 3-year mortality included ≥ moderate tricuspid regurgitation, ejection fraction less than 50%, and home oxygen use.

**Figure 4:**
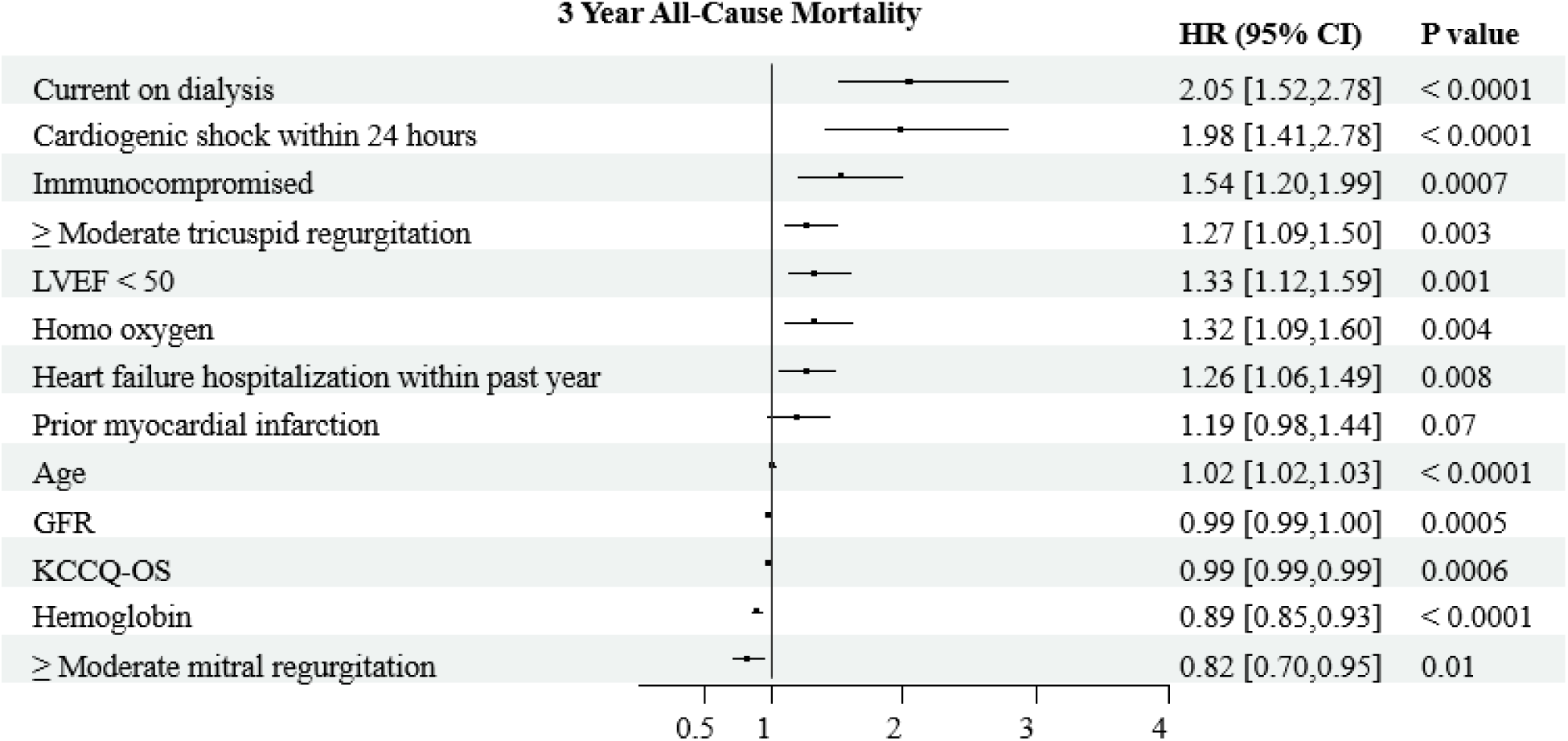
Predictors of 3-year all-cause mortality. Forest plot depicting significant predictors of all-cause mortality based on multivariable cox regression model. CI indicates confidence interval; GFR, glomerular filtration rate; HR, hazard ratio; KCCQ-OS, Kansas City Cardiomyopathy Questionnaire-overall summary; and LVEF, left ventricular ejection fractio

## Discussion

This large multicenter study is the first to present three-year clinical outcomes of patients undergoing transseptal MViV in the United States TVT Registry. Major findings from this analysis include: 1) Three-year mortality rates were relatively low in patients with low and intermediate STS scores (15.8% and 23.3% respectively), 2) Three-year mortality and stroke rates were higher in patients with high compared to low STS scores (44.5% vs. 15.8% and 11.4% vs. 7.6% respectively) 3) Elective procedure status was associated with lower 3-year mortality and similar stroke rates compared to non-elective procedure status and 4) Mitral valve reintervention rates at 3 years were similarly low (less than 4%) in all groups. These findings highlight the growing role of MViV in the longitudinal treatment of patients with mitral valve disease who require mitral valve replacement. In this study, durability of MViV was good at 3 years in all patient groups, with low rates of reintervention.

### Three-Year Survival in High-Risk Patients

Mortality rates during the first three years after transseptal MViV varied significantly depending on the severity of patient comorbidities. In patients with STS score greater than 8, mortality rate increased sharply to over 10% in the first 3 months despite a low procedural mortality rate of 0.6%, high device implant success rates, and low rates of procedural complications. Given the consistency of procedural results despite risk profile, it is likely that the higher mortality rate was driven by higher frequency of comorbidities, including more advanced age, kidney, lung disease, as well as more advanced heart failure with concomitant valvular heart disease. For example, high-risk patients had a five-fold higher rate of home oxygen use, seven-fold higher rate of cardiogenic shock, and seven-fold higher rate of renal failure requiring dialysis at the time of presentation, all of which were previously shown to be associated with increased mortality at 1 year of follow up.^2^ These important signs of advanced multi-organ dysfunction, which are more frequent in the high STS risk group, likely underly the contrasting survival following transseptal MViV. Of note, the mean STS predicted mortality in this group was 15.5% which further conveys the true risk beyond the definition of “greater than 8%”. Despite this higher mortality, the symptomatic response and quality of life improvements in patients who reached 1 year of follow-up were comparable to low and intermediate-risk groups, highlighting the fact that a large subset of high STS risk patients benefit greatly from this intervention. Similar findings have been demonstrated in transcatheter mitral valve replacement trials of high STS risk patients, where despite high 1-year mortality, large quality of life improvements were observed.^6,7^

### Three-Year Outcomes in Low-Risk and Intermediate-Risk Patients

Patients in the low-risk and intermediate-risk groups experienced high procedural success rates and low rates of stroke, reintervention, and mortality at 3 years of follow-up after transseptal MViV in the United States TVT Registry. Although the echocardiographic mean mitral prosthetic gradient was elevated post procedure, it remained stable at 1 year and rates of repeat intervention were low, suggesting that the elevated gradient was not deemed to be of clinical significance. It is plausible that some of these patients would be candidates for re-do cardiac surgery based on their STS risk scores if their MViV procedural result was unsatisfactory. Furthermore, mortality rates at 3 years were relatively low (15.8% and 23.3% predicted mortality at 3 years), accounting for the mean age of patients, with mean age 66.5 ± 12.7 years in the low-risk group and 73.8 ± 9.5 years in the intermediate-risk group. In a recent analysis of the intermediate-risk patients enrolled in the PARTNER 3 MViV Registry, there were no mortalities observed at 1 year of follow-up in 50 carefully selected patients, illustrating the minimally-invasive and successful nature of the MViV procedure when applied to patients without severe comorbidities.^9^ Importantly, even patients with a low STS risk score can be deemed “high-risk” for repeat cardiac surgery after heart team evaluation, often based on important variables including functional/ambulatory status and technical impediments such as porcelain aorta or hostile chest, among others. In the present study 47.5% of low STS risk and 57.4% of intermediate STS risk score group patients were deemed to be high-risk for cardiac surgery based on the site determination.

Medical management following successful MViV is an important aspect of longitudinal care. Prosthetic valve thrombosis is an important complication in MViV to be aware of, occurring in approximately 6% of patients during the first year.^9^ When detected early, prosthetic valve thrombosis can be treated successfully with anticoagulation therapy. To avoid prosthetic valve thrombosis, most patients undergoing MViV are treated with anticoagulation therapy. This was the case in the current overall study population, where 82.9% were prescribed anticoagulation at hospital discharge and 70.4% continued this therapy at one year follow-up.

### Importance of Surveillance and Timely Treatment

Several findings of the present study emphasize the importance of early identification of candidates for MViV to allow for prompt treatment. Patients with heart failure hospitalizations within the last year had higher 3-year mortality, suggesting that earlier identification and treatment may have an impact on prognosis. Patients undergoing urgent non-elective procedures had a nearly two-fold higher mortality at 3 years, further supporting this concept. Indeed, signs of advanced presentation such as cardiogenic shock on presentation and current hemodialysis use were some of the strongest independent predictors of 3-year mortality. Careful surveillance of patients with senescent mitral bioprostheses by a cardiovascular specialist with annual echocardiography is essential to identify prosthetic valve dysfunction promptly, which may lead to improved outcomes.

### Limitations

Study limitations include the observational design and the use of self-reported site data in the TVT Registry. Regular audits of the data are performed by the STS/ACC to maintain data accuracy. Although the majority of MViV procedures performed in the United States are captured in this registry, a minority of centers do not participate in the TVT Registry and their data are not included. Echocardiographic data beyond 1 year is not reported in the TVT Registry and thus is unavailable for this dataset.

### Conclusions

Three-year survival after MViV is favorable in low and intermediate STS score populations and significantly lower in high STS score patients treated in the United States. Non-elective procedures and patients with advanced heart failure presentation and multiorgan dysfunction had the highest 3-year mortality, emphasizing the importance of early identification and treatment of patients who may benefit from MViV. Reintervention rates at 3 years are low regardless of STS score.

## Data Availability

Data may be available upon reasonable request

## Non-standard Abbreviations and Acronyms

ACC: American College of Cardiology
CMS: Centers for Medicare and Medicaid Services
HR: hazard ratio
KCCQ: Kansas City Cardiomyopathy Questionnaire
LVOT: left ventricular outflow tract obstruction
MR: mitral regurgitation
MS: mitral stenosis
MViV: mitral valve-in-valve
NYHA: New York Heart Association
STS: Society of Thoracic Surgeons
TVT: Transcatheter Valve Therapy

## Acknowledgements

The authors would like to thank Angila Sewal, PhD, Edwards Lifesciences, for ensuring technical accuracy of the manuscript and for assisting with tables and figures for the manuscript, and Ke Xu, PhD, Edwards Lifesciences, for statistical support. They were not compensated for their contributions uniquely for this article.

## Supplemental Material

Tables S1-S8

